# Does the school feeding program have an impact on the development of human capital? Results from the Burkina Faso experience using the micro-simulation method

**DOI:** 10.1101/2025.06.05.25329100

**Authors:** Somwaoga Daniel Ouedraogo, Ella W. R. Compaore, Gustave Adrien Nebie, Thierry S. Z. C. Coulibaly, Johanne Desormeaux, Ousmane Ouedraogo, Saidou Diallo, Inoussa KY, Mamoudou H. Dicko

## Abstract

School feeding program is one of the world’s most popular interventions for school-age children. Under-investment in this age group is a major obstacle to human capital development. This study aims to provide evidence on the impact of school meals on student survival, nutrition and education. This study is based on the method of dynamic micro-simulation of the behavior of schoolchildren consuming school canteen meals. The statistical software R was used to process both the individual data and to write the model. A cohort of 1,000 beneficiary schoolchildren was used to observe the impact of this canteen during the primary cycle. Simulation at 100% coverage of the school year shows a cost USD 18,489 per year and per beneficiary, and over the six years of the primary cycle: i) saves three lives out of every 1,000 pupils, ii) gives each pupil around 3 months more life, iii) saves 121 pupils from dropping out of school, and iv) reduces the level of stunted growth from 18.5% to 9.8%, i.e. a reduction of 8.7% in six years. The results indicate the higher the level of school feeding coverage, the greater its impact on human capital development in Burkina Faso.

## Introduction

School feeding programs are a long-standing global public health standard, focused on improving health and education outcomes for socio-economically disadvantaged students (1, 2). According to the World Bank’s State of Social Safety Nets 2018, school feeding is not only the largest safety net in the world but also the most widespread, considering the number of countries that have adopted it and the number of beneficiaries (3). This underlines the fact that not only has school feeding become the main intervention for children in school, it is also the most widespread safety net in the world, whatever the category or age group of beneficiaries.

In fact, 418 million children, or 41% of children enrolled in primary school, will benefit from school feeding programs worldwide in 2022 (4).

The objectives of school feeding programs can include the alleviation of short-term hunger (5, 6) and the improvement of micronutrient status, growth (5), cognition (7, 8) and school performance (1, 7, 9, 10).

Stunting as an indicator of nutritional status is that, at the microeconomic level, it has been shown that a 1% loss in adult height due to stunting in childhood is equivalent to a 1.4% loss in individual productivity (11). Stunting is worrying because the resulting loss of height and intelligence is permanent. Moreover, it is intergenerational and therefore perpetuates the loss of productivity for individuals in subsequent generations. Consequently, investment in improved nutrition to put an end to stunted growth can be translated into investment in human capital and intergenerational transfers (12).

A series of analyses published since 2017 have highlighted the need to invest in children’s health, nutrition and education throughout the first 8,000 days of their lives if they are to grow up to fulfil their potential as adults (13-15). Schools are the key to implementing this health, nutrition and education interventions, and are therefore essential platforms for the development of human capital, which contributes more than 70% of the wealth of high-income countries is attributed to human capital, compared with 40% in low-income countries (16-18).

Investment in human capital, which is the wealth of nations, is multidimensional and complementary, and involves three components: survival, health and nutrition, and education. (19). While the development of human capital depends on quality education, good health and nutrition are also necessary for school-age children and adolescents to participate and learn in school and as adults in order to be productive. It is during this age group (5 to 19 years) that this population group requires particular attention from the education and health sectors as they undergo physical, emotional and cognitive changes (13, 17, 18, 20).

In Burkina Faso, the history of the school canteen is closely linked to that of the introduction of schooling by the colonists in the 1900s. As the school system progressed, so did its operation. In order to guarantee these children better study conditions, the colonial administration opened boarding schools and set up the first school canteen, which operated solely on the basis of food collected by pupils’ parents and/or local produce (21, 22).

During the post-colonial period, on 16th August 1960, the government of Upper Volta signed an agreement with Catholic Relief Services (CRS), through the Cathwell project, for food assistance in the fields of education and health. Following this agreement, CRS offered, from 1962 until 1988, to take full responsibility for feeding all schoolchildren following a serious famine in the Sahel region of West Africa. With the signing of this agreement between the Government of Burkina Faso and the CRS, the endogenous school canteen will become an assisted school canteen (21).

However, for financial and strategic reasons, the NGO decided to refocus its intervention. This reorientation resulted in the subtraction of large cities from aid, in order to focus on areas with very low school enrolment rates and food insecurity. Management of the school canteen program was then transferred to the Ministry of Education. CRS now intervenes in rural areas where school enrolment rates are very low and food insecurity is a reality (23).

Burkinabe authorities, aware that the canteen has a very positive impact on the quality and efficiency of education, set up a school canteen service in 1988, attached to the Department for the Allocation of Specific Resources to Educational Structures (DAMSSE) at the Ministry of Education.

Since 2011, in response to food insecurity in many localities, the government has been automatically providing all schools with canteens. For reasons of insufficient funding, the state allocation only covers around 3 months of classes out of the 8 months. School allocations must respect the following composition and ratios: cereals (rice): 67%; legumes (beans): 16%; oilseeds (oil): 17%. These foodstuffs are used to prepare school meals for pupils (24).

Furthermore, in 2023, the Burkina Faso government spent USD 31.12 million, including USD 30.13 million for primary schools (3,722,119 pupils) and USD 982,508.2 for pre-school (121,371 children) (25).The aim of this study is to simulate the impact of the school feeding program on human capital development in Burkina Faso. The mechanism for transmitting the impact of the school feeding program on individuals is therefore essentially through improved nutritional status, survival, reduced absenteeism and improved learning, all of which are fundamental components of human capital development.

## 1. Methodology

The study on the impact of school feeding on the development of human capital was carried out using the micro-simulation technique. It used the free statistical software R to process both the individual data and to write the model. The R packages heemod, chron, scales and ggplot2 were used. All calculations were performed using Microsoft Excel 2019. Micro-simulation is an investigative method based on the individual reproduction of the behaviours studied, using a representative sample. This study involved comparing canteen costs with the results of studies in other countries in order to highlight the impact on pupils’ nutritional status (26), school retention and pupils’ lives (2, 27).

In this case, the analysis consisted in defining several coverage scenarios (five scenarios) for the school canteen in public elementary school, compared with the current level (3/8 months), and analyzing the impact of each level of coverage on: nutritional status (“stunting”)(28), school drop-out (29), quality of life (QALY) (30, 31) and the number life saved for schoolchildren.

QALYs (Quality adjusted life years), simultaneously take into account the impact of the school canteen on the duration and quality of life of students. To calculate QALYs, life-years gained are weighted by a health-related quality-of-life factor ranging from 0 (death) to 1 (best possible health).

The QALY is then used to calculate the number of additional years of healthy life gained, depending on the level of coverage of the school canteen throughout the simulation.

In order to compare school canteen coverage scenarios in terms of cost and impact on pupils’ lives, and to provide more information on the “profitability” of a scenario, the Incremental Cost Effectiveness Ratio (ICER) will be calculated.

This is the ratio between the difference in cost of two strategies and their difference in effectiveness. The ICER will thus enable us to determine the additional cost generated per unit of effectiveness obtained on: stunting, school drop-out, number of lives saved and number of additional years of life gained. This effectiveness can be measured and given in life-years gained, or in life-years gained weighted by quality of life, in QALYs.

### 1.1. Data sources and quality

The data used in this study come from official documents of the Ministry of National Education, Literacy and the Promotion of National Languages (MENAPLN) for the 2022/2023 school year. These are mainly the statistical yearbook (29) and the interministerial decree on the resources transferred to the communes to support education (25). The target population for this study is first grade (or “first Grade”) pupils attending public primary schools in Burkina Faso, which represent a total population of 393,973 schoolchildren (25).

### 1.2. Basic assumptions based on various studies

The following impact chain **(Figure 1)** was used as the impact chain of the school canteen on the development of human capital.

**Figure 1:**
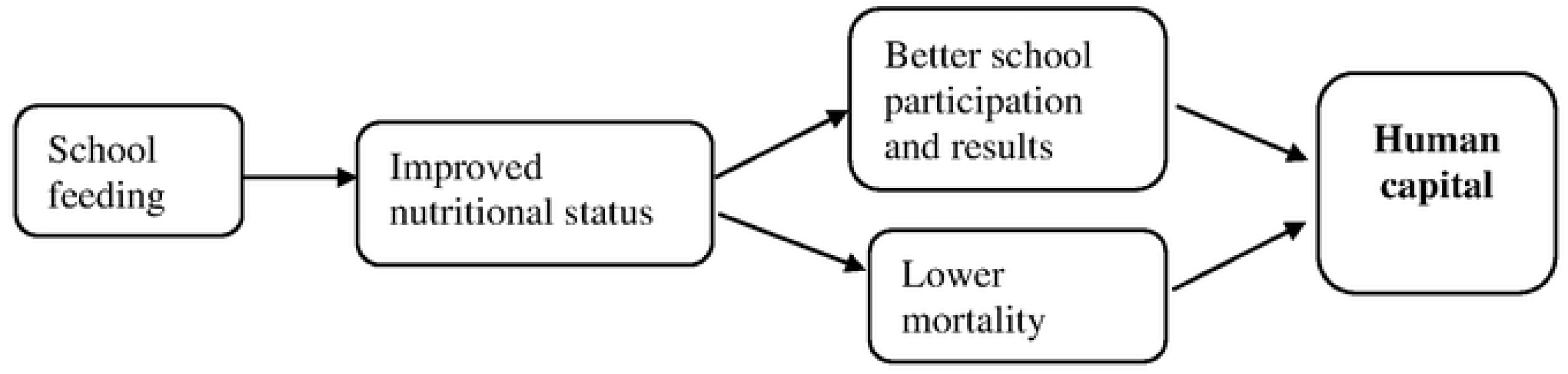
Hypothesis of the irnpact way of school feeding prograrn on hurnan capital development

Setting up a school meals can have the effect of improving the nutritional status of beneficiary pupils (32-34). Improved nutritional status has a positive impact on: better school results (33, 35, 36) (links between nutrition and school performance) and better health (37) (links between nutrition and health), which should result in lower mortality (1, 38). Better school performance and good health are essential for the development of human capital (32-34). In order to design the model, the following hypotheses were developed:

- school canteens have an impact on the nutritional status of beneficiary pupils (5, 32, 39);
- Pupils attending schools without functional school canteens have an incidence of malnutrition 2.5 times higher than those with access to canteens (32);
- no-malnourished pupils have a school attendance rate 4 times higher than those who are malnourished (7, 9, 40);
- Death rates for malnourished children are 5 times higher than for non-malnourished children (41);
- all schools are covered by school canteens, but for only 3 months, compared with 8 months of classes over the nine-month school year. We therefore assume that only 3/8 of pupils are covered by school canteens (24);
- malnutrition is approximated by stunting, the rate of which is currently around 22% in Burkina Faso (42);
- the canteens cover all primary schools with a beneficiary age range of between 6 and 14 years (24, 35);
- it’s assumed that there is no grade repetition for the pupils.

### 1.3. Projection assumptions

The following assumptions have been made in the projection levels of coverage by the school canteen with 2024 as the base year:

- the projection was made over 6 years, corresponding to the normal length of the primary cycle for a pupil who does not repeat a class;
- we have set ourselves a target of increasing school canteen coverage, with current coverage of 3 months out of the 8 months of school (3/8): TIC=3/8;
- in the baseline scenario, TIC = 0, i.e. it is assumed that there is no improvement in school canteen coverage compared with the current situation;
- in the other scenarios, it is up to the user to choose a rate of increase in school canteen coverage.

The model was used to develop six potential scenarios based on the extent to which the school canteen covers school periods.

1. 3 months called **‘Base’**. In this strategy, only 3 of the 8 months of classes are covered by the canteen. The reference scenario corresponds to the current school canteen situation in Burkina Faso;
2. 4 months of classes, referred to as **‘scenario 1’**.
3. 5 months of the 8 months of classes, referred to as **‘scenario 2’**.
4. 6 months referred to as **‘scenario 3’**.
5. 7 months out of the 8 months of the school year known as **‘scenario 4’**.
6. 8 months of the school year (100%) referred to as **‘scenario 5’**. In this strategy, the canteen is operational for all 8 months of the school year.

### 1.4. Input parameters

January 1, 2024 is the starting year of the simulation (D<- dates) with a sample of 1000 pupils (n.i <- 1000) over a time horizon of six years (n.t <- 6); i.e. during the entire primary cycle of the 1000 pupils and on the assumption that there will be no cases of repetition. All the children were born on 1 January 2018, so were 6 years old on January 1, 2024. The school canteen coverage rate (CCR) in 2024 is 3 months on average out of 8 months of classes (CCR <- 3/8 = 0.375). The model was developed on the basis of the current annual unit cost of the school canteen, which is estimated at USD 27.87 (canteen cost <- 27.87) (25). In order to calculate the impact of the school canteen on pupils’ lives, the “**Quality Adjusted Life Years** (QALYs) (30, 43, 44)” were calculated, corresponding to the years of life gained by each pupil by consuming the school canteen meals with the hypotheses. To calculate the QALYs, the years of life gained are weighted by a health-related quality of life factor ranging from 0 for death to 1 for a year in good health.

### 1.5. Possible states

For nutritional status (v.nut <- c(‘MNUT’, ‘BNUT’)), two cases are possible: malnourished (MNUT) or well nourished (BNUT). With regard to the impact on pupils‘lives, two possible outcomes called states of life (v.dead <- c(’VI’, “MR”)) are expected with “VI” as alive and “MR” as dead. In terms of education, the impact is measured in terms of school dropouts. The result will then be v.edu <- c(‘PRI’, ‘OPRI’) with ‘PRI’ as staying in school and ‘OPRI’ as dropping out.

### 1.6. Transition probabilities by cycle

#### 1.6.1. Nutritional transition: stunting

A ratio of 2.5 (rrCS <- 2.5) between the malnutrition rate of pupils who do not benefit from school canteens and those who do (33, 34) was introduced on the basis of a stunting proxy (TRC <- 0.22) for school-age children in Burkina Faso of 22%. On this basis, the projection was made using the following evidences:

- prevalence of malnutrition in children with access to school canteen meals (TMC) (TMC <- TRC/(TCC+rrCS-rrCS*TCC) (32-34);
- prevalence of malnutrition in children without access to school canteen meals (TMNC <- rrCS*TMC) (32-34);
- probability of a child with access to school canteens becoming malnourished (p.TMC <- rate_to_prob (r=TMC, per = 1));
- probability of a child without access to school canteens becoming malnourished (p.TMNC <- rate_to_prob(r=TMNC, per = 1)).

#### 1.6.2. School transitions

Drop-out is the ratio of the number of pupils who leave the school cycle during the school year for any reason to the total number of pupils enrolled during that same school year. It can be calculated at primary, post-primary, secondary or tertiary level (29). It is assumed that there is no repetition.

The ratio of school attendance rates (rrFS) between malnourished and non-malnourished pupils varies between 2 and 6 (7, 36, 45), depending on the study. In the lack of data, we have taken the average of the two extremes, which is 4 for Burkina Faso in this simulation (rrFS <- 4). Primary completion rate 2022 in Burkina Faso over 6 years was 54% (TFS <- 0.54) (46). The completion rate for malnourished children was calculated in the sample using the formula: TFM <- TFS/(TRC+ (1-TRC)*rrFS) and the completion rate for non-malnourished children using: TFNM <- rrFS*TFM. For these calculations, the probability of completion for a malnourished child is: p.TFM <- rate_to_prob (r=TFM, per = 1) and that for a non-malnourished child is: p.TFNM <- rate_to_prob(r=TFNM, per = 1). Thus, the probability of completion for a malnourished child over 6 years is: p.TFMA <- rescale_prob(p=p.TFM, to = 6) and that over the same period for a non-malnourished child is: p.TFNMA <- rescale_prob(p=p.TFNM, to = 6).

#### 1.6.3. Dead transition

The annual mortality rate (AMR) for children in the 5-14 age group (%) is 6.9% [43]. It is estimated that 40% of total child mortality is due to malnutrition (NMR <- 0.40). To do this, the mortality rate for non-malnourished children over 10 years is: TMENM <- TMGE*(1- TMN) and that for non-malnourished children is: TMEM <- rrMort*TMENM. The probability of death over the period 5 to 14 years for a pupil is: p.MENM <- rate_to_prob(r=TMENM, per = 1). The annual probability of death for a non-malnourished child is equal to: p.MENMA <- rescale_prob(p=p.MENM, to = 1/10). The mortality rate ratio between malnourished and non-malnourished children is: rrMort <- 5. The annual probability of death for a malnourished child is calculated using the formula: p.MEMA <- rescale_prob(p=p.MEM, to = 1/10).

### 1.7. Ethical considerations

The study was officially approved by the Burkina Faso Health Research Ethics Committee (CERS) with reference N 2022_33_/MS/MESRSI/CERS of 14-02-202. In addition to the approval of the Ethics Committee, the study was authorised (Ref. N 000115/MENAPLN/SG/DAMSSE of 21-01-2022) by the Ministry in charge of National Education.

## 2. Results

A simulation of a cohort of 1,000 pupils is used to project the model over the entire primary cycle, i.e. six years, simulating school canteen coverage rates in relation to current levels. To do this, five other coverage scenarios were investigated in relation to the current level of school canteen coverage, in order to analyse the impact of each level of coverage on the lives of pupils (number of lives saved, quality of life gained and unit cost), education (dropping out of school) and the nutritional status of pupils (stunted growth). The simulation also provided the cost (unit, annual) of the school feeding program according to the level of coverage, as well as the costs of the program. At the end of the 6 years and according to the scenarios and modelling hypotheses explained in the previous section, the following simulation results were obtained. For all the results obtained, we note a substantial homogeneity based on the variation in the level of coverage of the school feeding. For example, the scale of the gains in terms of lives (number saved, quality), education and nutrition depended on how long the school canteen was in operation.

### 2.1. Number of lives saved according to coverage rate

**Table 1** below shows the evolution of the number of pupils whose lives were saved according to the different scenarios over a 6-year primary cycle according to the level of coverage of the school canteen over the 8 months of the school year. The impact on pupils’ lives is significantly felt from 50% coverage onwards, with one life saved.

**Table 1:**
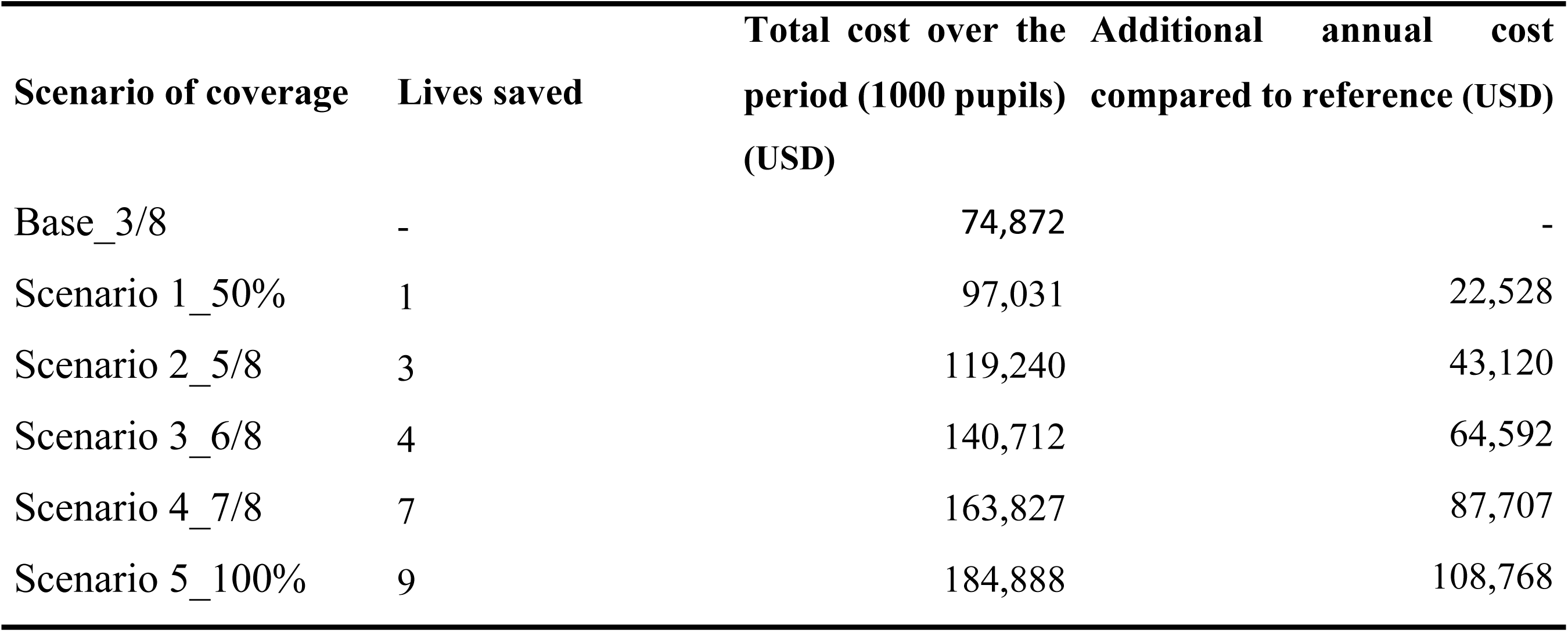
Variation in the number of lives saved as a function of school meals costs over the 6-year projection period and according to the level of coverage for a sample of 1,000 pupils from 2024 to 2030.

From a coverage rate of 50% (scenario 1), it can be seen that the school canteen saves lives, with the number of lives saved increasing according to the level of coverage. Coverage of the school feeding program for 5 months saves 3 lives among beneficiaries during the primary cycle. The scenario in which the canteen is operational for five or eight months saves two lives each. Finally, in the most favourable scenario, 100% coverage or ‘Scenario 5’, the number of lives saved is nine pupils per thousand at the end of the 6th year of primary school. As a result, “scenario 5” would save the lives of nine (9) schoolchildren over the entire cycle, at an additional cost of USD 106,984.92.

These results show that operating the school canteen for 5 months, at a cost of USD 117,285.24 per year, would save as many lives (2 lives) as operating the canteen for 6 months (USD 138,405.24) or 7 months (USD 161,140.98).

This table also gives the total costs of school meals for the simulation sample of 1,000 pupils. These costs naturally vary according to the level of coverage. For example, these costs rise from USD 76,120 over the entire primary cycle (first grade to sixth class), 6 years, for a canteen operating for 3 months, to USD 184,888 over 6 years, or USD 30,815 per year for the school canteen to operate for the entire school year (9 months). The additional costs according to the simulated level of coverage compared with current coverage were calculated.

In relation to the 393,973 pupils in first grade classes in public primary schools during the 2022/2023 school year in Burkina Faso (25), the school canteen with 100% coverage during the school year would save the lives of 1,182 pupils during the cycle. To achieve this, the government would have to invest USD 72,840,880 million over the six years of the primary cycle, or USD 12,140,147 million per year.

### 2.2. Impact on quality of life (QALY)

In this study, the number of years of life gained or QALY (Quality Adjusted Life Years) per pupil thanks to the consumption of the school canteen meal was calculated per scenario. The QALY was estimated according to the level of canteen coverage.

The baseline scenario, which corresponds to the current level of coverage of the school canteen (3 months of operation out of 8 months of classes), at USD 76.12 during the 6-year primary cycle, or USD **12.70** per child per year, gives a QALY level of 5.33, i.e. a child will live in good health for 5 years and 4 months out of the 6 years of primary school. (**Table 2)**.

**Table 2:** Year equivalents and additional life-years saved per pupil by level of school canteen coverage and additional annual costs over the 6-year projection period by level of coverage from 2024 to 2030.

| Scenario of coverage | QALY (tQ<sub>hat</sub>) | Additional QALY | Average cost per pupil over the period (tCant<sub>hat</sub>) (USD) | Average annual unit cost (USD) |
| --- | --- | --- | --- | --- |
| Base_3/8 | 5.33 | 0 | 76.12 | **12.70** |
| Scenario 1_50% | 5.38 | 0,06 | 98.50 | **16.64** |
| Scenario 2_5/8 | 5.42 | 0,10 | 119.24 | **19.87** |
| Scenario 3_6/8 | 5.49 | 0,16 | 140.71 | **23.45** |
| Scenario 4_7/8 | 5.54 | 0,22 | 163.83 | **27.31** |
| Scenario 5_100% | 5.59 | 1,27 | 184.90 | **30.82** |

A simulation to 100% coverage gives an impact on the quality of life of pupils of 5.59 years. The results show an improvement in the quality of life of pupils receiving school canteen meals as a function of the level of coverage.

The improvement in quality of life as a function of the school canteen coverage scenario makes it possible to calculate the number of additional years of life gained as a function of the current level of coverage (3/8 months).

This shows that a 50% coverage (scenario 2) of the school year by the school canteen, with a unit cost of USD 98.65 per pupil during the primary cycle, enables beneficiaries to gain 0.06 QALYs compared with the baseline scenario. This positive impact increases with the level of canteen coverage, with QALYs of 0.1 (Scenario 2), 0.16 (Scenario 3) and 0.22 (Scenario 4) respectively (**Figure 2**).

**Figure 2:**
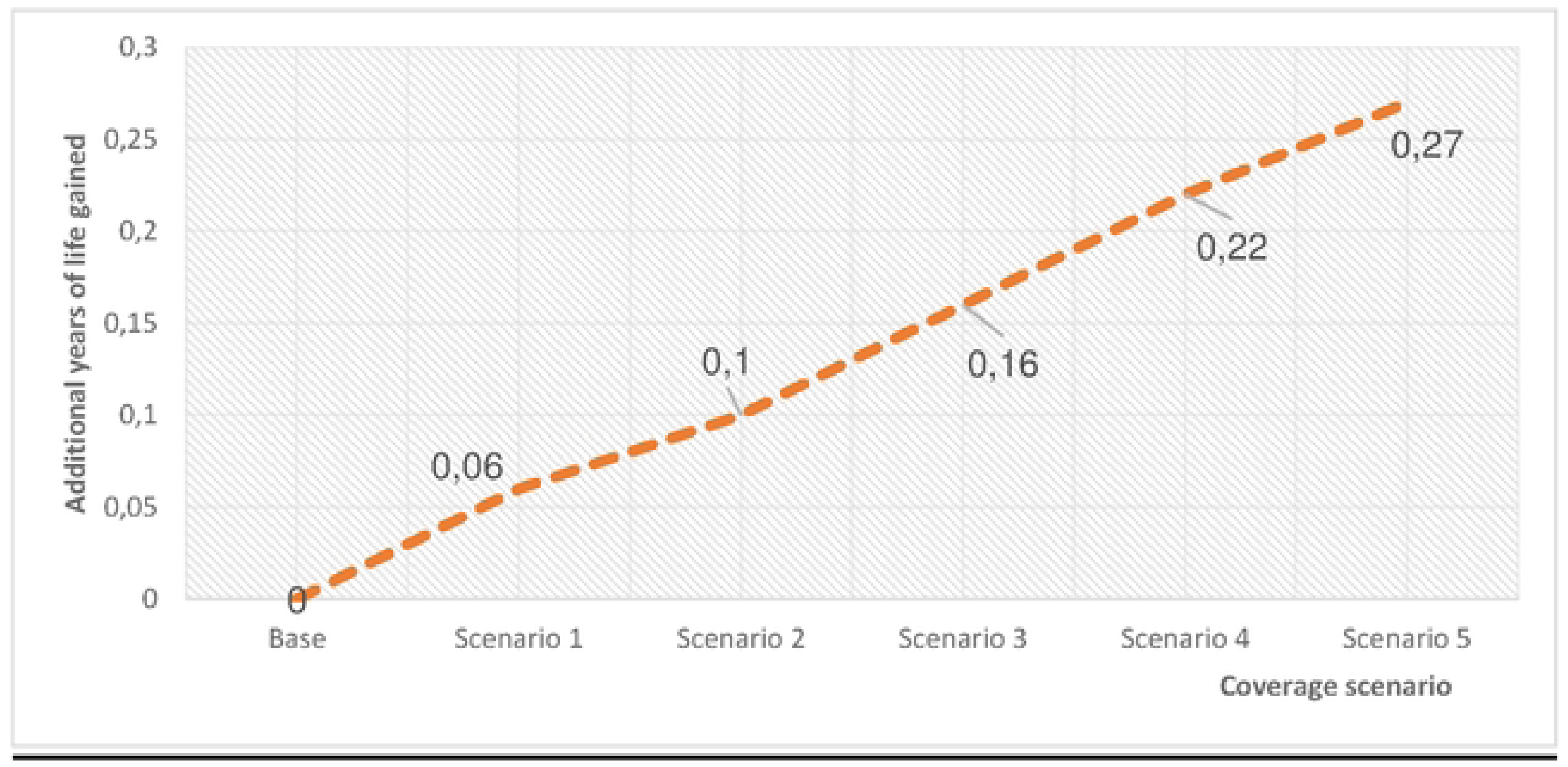
Nu1nber of additional years of life saved for pupils benefiting fro1n school canteen n1eals over 6 years, according to the level of coverage co1npared with the Baseline situation.

A 100% coverage of the school year (Scenario 5) at a cost of USD 184.90 **per pupil, or an annual cost of** USD **30.82,** gives a QALY of 0.27 additional years of healthy life after six years. In the latter scenario, a child will live on average 3 months longer in perfect health, compared with the baseline scenario.

### 2.3. Unit costs of quality of life gained

The data were used to calculate the Incremental Cost Effectiveness Ratio (ICER) (47) (**Table 3)**. It is used to determine the additional cost generated per unit of effectiveness (QALY) obtained or number of life years gained. **Table 3** below shows the variations between scenarios in the costs and impacts obtained. ICER is a ratio of cost per life-year gained or cost per life-year gained weighted by quality of life.

**Table 3:**
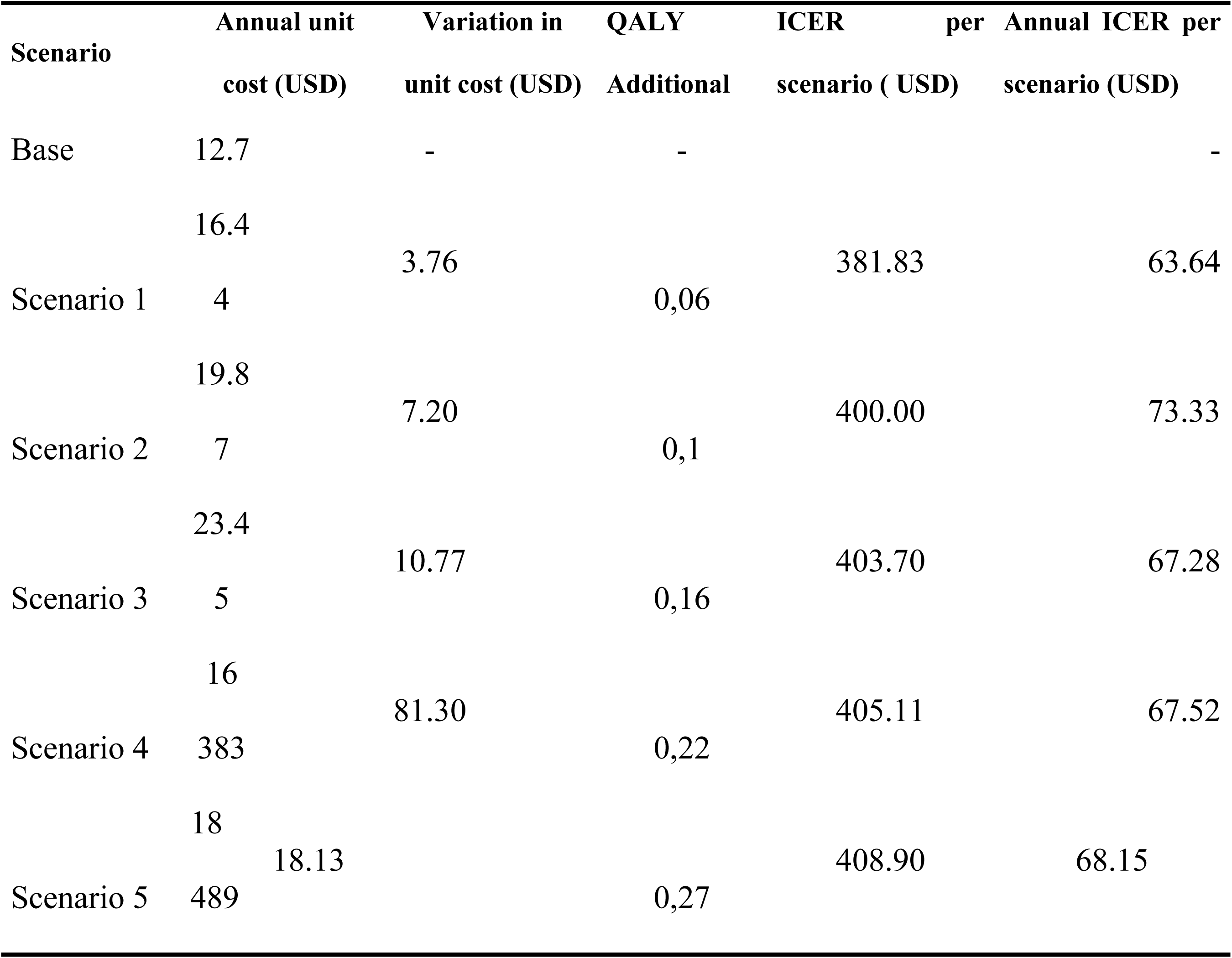
Variation of financial costs of life-years saved per pupil according to school canteen coverage levels over the 6-year projection period from 2024 to 2030.

The table compares the difference in costs between the coverage scenarios (Scenario 1) and the reference scenario (Base), as well as the number of additional years of life (additional QALYs) gained. It can be seen that providing 50% coverage for children (Scenario 1) results in an additional unit cost of USD 3.76 per child per year, and a gain of 0.06 additional years of life after six years, compared with the baseline scenario. The Differential Cost/Result Ratio (DCRR) or ICER for this scenario is USD 63.64 (ratio USD 3.76 to 0.06) per year, or USD 381.83 over 6 years.

A 100% coverage (Scenario 5) of the school year by the canteen, compared with 50% coverage (Scenario 1), costs an additional USD 18.13 per child and increases the QALY by 0.27 years. This level of coverage gives an annual ICER of USD 68.15 (ratio USD 18.13 to 0.27). Thus, over the six years of the intervention, USD 408.90 need to be spent to obtain 1.3 years of life.

### 2.4. School drop-out

The dropout rate in this study is the number of students in a cohort enrolled in a given year of study who drop out the following school year. This number is determined for intermediate classes, i.e. those that do not end with a diploma. Ideally, the number of dropouts should be as low as possible. A high drop-out rate is a sign of internal efficiency problems in education systems. In Burkina Faso in 2023, the average dropout rate will be 20.08%, with 21.1% for first grade and 27.6% for fifth class (29) in school. During this period, the completion rate for this cycle was 54.6% (46). **Table 4** shows the impact of the school canteen on primary school drop-out. For 1,000 children enrolled in primary school, the scenarios show how the school canteen, depending on the level of coverage, contributes to reducing the number of dropouts from first grade to sixth class.

**Table 4:** School drop-out variation and reduction gaps among 1000 schoolchildren according to the level of school canteen coverage over a 6-year projection from 2024 to 2030.

| **Scenario** | **Total cost of canteen over the period (USD)** | **Additional cost compared to reference (USD)** | **School drop-out** | **Number of dropouts avoided compared to baseline** |
| --- | --- | --- | --- | --- |
| Base_3/8 | 76,120 | - | 423 | - |
| Scenario 1_50% | 98,648 | 22,528 | 388 | 35 |
| Scenario 2_5/8 | 119,240 | 43,120 | 383 | 40 |
| Scenario 3_6/8 | 140,712 | 64,592 | 362 | 61 |
| Scenario 4_7/8 | 163,827 | 87,707 | 325 | 98 |
| Scenario 5_100% | 184,888 | 108,768 | 302 | 121 |

In the basic scenario corresponding to the current level of canteen coverage, which is 3 out of 8 months, on the basis of 1 000 children enrolled in first grade for the 2023/2024 school year, around 423 children will have dropped out by the time they reach sixth class in the 2029/2030 school year.

In the case of 100% coverage (scenario 5), the number of pupils dropping out of school during the cycle is lower than in the baseline scenario. Out of 1,000 children enrolling in first grade class in 2024 who benefit from canteen meals throughout the school year and the entire primary cycle, 698 pupils will reach sixth class, i.e. 302 dropouts. The number of pupils who will have dropped out throughout the primary cycle compared with the baseline scenario will fall from 423 to 302. Thus, with an additional cost of USD 108,768 for the canteen compared with the baseline scenario (USD 76,120), 121 pupils are saved from dropping out.

On a national scale, out of the 393,973 pupils in first grade classes in Burkina Faso in 2023 with a canteen that operates for 3 months of the school year, the simulation shows that 166,650 pupils will drop out of classes during the primary cycle. On the other hand, with a canteen operating throughout the school year, by the end of the cycle 118,978 pupils will have dropped out by the 2027/2028 school year. A functioning canteen would therefore prevent 47,670 pupils from dropping out.

### 2.5. Impact on stunting reduction

Analysis of changes in the prevalence of stunted pupils according to the different scenarios shows the following results (**Figure 3**). The results of the different scenarios show a reduction in stunting according to the level of investment in the school canteen.

**Figure 3:**
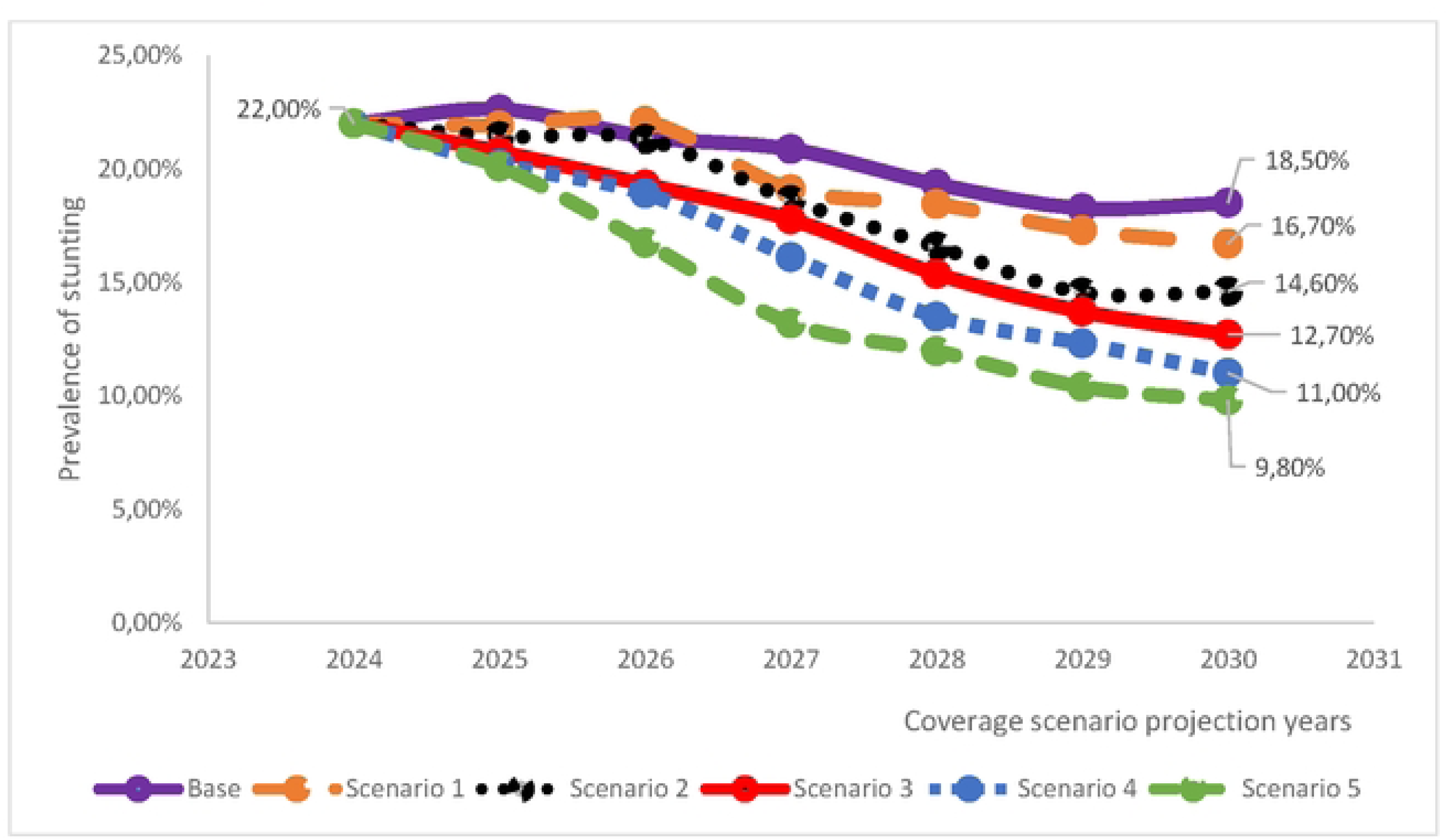
Prevalence of stunting among 1.000 children receiving school meals in primary school. by scenario of coverage

In the reference scenario, the prevalence of stunting among pupils was 18.5%. In the 50% coverage scenario, the incidence of stunting falls to 16.7% over six years, a reduction of 1.8% compared with the baseline (**Table 5)**.

**Table 5:** Variation of stunting prevalences and reduction gaps among 1000 schoolchildren according to the level of school canteen coverage over a 6-year projection from 2024 to 2030.

| Scenario | Stunting effect status (%) | Growth delay avoided according to the level of cover in relation to the base (%) |
| --- | --- | --- |
| Base_3/8 | 18.5 | - |
| Scenario 1_50% | 16.7 | 1.8 |
| Scenario 2_5/8 | 14.6 | 3.9 |
| Scenario 3_6/8 | 12.7 | 5.8 |
| Scenario 4_7/8 | 11.0 | 7.5 |
| Scenario 5_100% | 9.8 | 8.7 |

In the scenario of 100% coverage of pupils by the school canteen throughout the school year, stunting would fall to 9.8% over six years. This would reduce the prevalence of stunting among pupils by 8.7% compared with current coverage.

## 3. Discussion

To the best of our knowledge, this is the first study that explicitly examine from a holistic perspective the simultaneous impact of a national school meals program on human capital in Burkina Faso.

In Burkina Faso, the estimated unit cost of the canteen over the three months of operation is 11 US dollars per child which represent a daily unit cost is 0.18 US dollars per meal. The state provision covers 3 months of the school year, with 20 days of classes per month, i.e. a total of 60 days out of the approximately 175 days of classes during the school year. This annual unit cost is still far from the 40 to 50 US dollars per year per beneficiary estimated by the World Bank in 2012 (48).

### 3.1. Number of lives saved according to coverage rate

The results of the coverage simulations show that 100% coverage of schools during the school year would save the lives of three (3) pupils over the entire primary school cycle out of a sample of 1,000 pupils enrolled. This implies additional costs of USD 108,768 over the cycle compared to the costs of the current level of coverage, which amounts to USD 76,120 over the cycle. Thus, to save three (3) lives over 6 years, it would be necessary to invest an additional USD 18.13 each year for each pupil, on top of the annual unit cost of approximately USD 12.70. These results imply that of the 393,973 pupils enrolled in first grade primary cycle in Burkina Faso for the 2022/2023 school year, approximately 1,182 pupils will have their lives saved at the end of the Year 6 primary cycle (**sixth class**) if the school canteen covers the entire school year. The results of similar studies are not available for comparison. This is one of the few studies to analyse the impact of the school canteen on pupil mortality.

Undernutrition and micronutrient deficiencies can adversely affect physical of child health and development. Effects on physical health may include being underweight, stunted growth, lowered immunity, and mortality (6). Studies have shown that the school feeding programme has a significant effect, improving illness of beneficiary students (49, 50). By making a positive contribution to improving pupils’ illness, the school feeding saves lives among beneficiary. Therefore, school feeding programs have been a longstanding global public health standard, focused on improving health and education outcomes for students experiencing socioeconomic disadvantage (51).

### 3.2. Number of life years gained (QALYs)

The research also calculated the number of years of life that beneficiary pupils gain by eating school canteen meals.

The number of QALYs gained as a result of the improvement in the quality of life due to the school canteen increases according to the level of coverage during school periods. A pupil who benefits from school canteen meals throughout his or her primary education would gain 0.27 years compared with the baseline scenario, or around 3 months of life. These results confirm that a varied and balanced school diet helps to meet the nutritional needs of pupils, promoting good health (51).

Indeed, it has been shown that when school feeding program are designed with a nutritional objective in mind, they can provide around 30-40% of the internationally recommended daily allowance for school-age children (5).

The impact of the school feeding program on the nutritional status of pupils will have an effect on their morbidity (52). Reducing morbidity will lead to more years of healthful life. This will enable pupils who benefit from the school feeding service to have more years of life than those who do not benefit from it.

The gains in QALYs are therefore substantial, but it is important to analyse at what cost.

### 3.3. Dropping-out of school

The world’s population is healthier and better educated than ever before. In 1980, only 5 out of 10 children of primary school age were enrolled in school in low-income countries. By 2015, this figure had risen to 8 out of 10 (19). In Burkina Faso, the net enrolment rate will be 61.5% in 2023. This net enrolment rate is marked by a dropout rate of around 20.08% in 2023 (29). The high drop-out rate in Burkina Faso could be explained by the difficult security situation, which has led to the closure of several schools. According to UNICEF, one in four schools in Burkina Faso, i.e. 6,149, was closed due to persistent insecurity in certain parts of the country at the end of the 2022-2023 school year, leaving around one million children out of school and exposed to numerous threats (53). Other factors include parental ignorance and poverty, early marriage, teenage pregnancy and hunger (54). All these factors, which have a negative impact on the school completion rate, prevent the country from honouring its national and international commitments. To this end, the country has focused on several approaches, including school feeding, aimed at achieving the objectives of the international commitments Burkina Faso has ratified, including the Sustainable Development Goals (SDGs), the Continental Strategy for Education 2016-2025, the African Union’s Agenda 2063 and the Incheon Declaration. Also, national commitments to strengthen the education system, including: i) the Strategic Plan for the Development of Basic and Secondary Education (SPDBSE), ii) the 2021-2030 language policy, iii) the 2017-2030 Education and Training Sector Plan (ETSP). The ultimate aim of these commitments is to increase human capital through the acquisition of knowledge, skills and competencies. In view of the challenges involved, the government has adopted the Presidential Initiative: ‘Ensure that every school-age child has at least one balanced meal a day’ (55) and the National School Food and Nutrition Strategy (NSFNS) for Burkina Faso 2021-2025 (56). The Government’s policy will doubtless have a positive impact on the nutritional status of pupils. This impact will be reflected in school results. Simulation results show that the school canteen helps to reduce the number of children dropping out of school. The school feeding program ensures that every child has at least one meal a day (8, 37), means that pupils no longer have to walk long distances at midday to get something to eat, and gives them more time to revise their lessons (6, 35, 57). As a result, school attendance is improved (10, 35), which reduces the drop-out rate (58). The positive impact on school drop-out has also been demonstrated in studies carried out in the Jigjiga area of Somalia (59), in Osun State in Abuja, Nigeria (60, 61) and in Morocco (62). Nutrition and health programs in schools have been shown to influence drop-out rates (10).

Human capital investment theory attributes differences in individual earnings to differences in talent, family background, wealth and other assets (15). On average, for an individual, an additional year of schooling increases overall hourly earnings by 9% (63). These gains are significant in low- and middle-income countries, particularly for women (17).

Reducing school drop-out rates is not synonymous with increasing cognitive skills. A study in Sweden shows that school feeding programs have an effect on academic results, but not on cognitive test scores (64). The absence of effects might reflect that the school feeding program was introduced at ages beyond the critical periods for the development of cognitive skills (65). Another possible explanation is that school canteen meals increased human capital accumulation by making students more attentive and increasing their concentration time e at school. If meals have made it easier for students to learn, it’s possible that this has increased their level of education, even if cognitive abilities have not been affected (64).

### 3.4. Impact on stunting reduction

The school feeding program clearly has positive effects on the nutritional status of pupils. The results of the scenarios show a reduction in the prevalence of stunting according to the rate of coverage of the school canteen. In fact, the prevalence of stunting falls from 18.5% to 9.8% when the school canteen is extended from 3 months to 8 months, which is the length of time classes are actually open in Burkina Faso.

Studies carried out to compare changes in the prevalence of stunting among pupils receiving school meals and those who do not show that the canteen has a positive impact on the growth of pupils. In Kano, Nigeria, 91.8% of pupils who benefited from the school meals program had normal growth compared with 81.2% of pupils who did not (66). This justifies the fact that school meals meet the micronutrient requirements for proper physical growth. In the same trend, in 2018, pupils in schools not participating in the school meals program had higher rates of stunted growth than those benefiting from the program (39, 67). These results support the findings of a related study conducted over a two-year period in Lao PDR in 2011, which had already identified a positive impact of the school feeding program on the nutritional status of pupils (68). Similarly, in 2019, research into a school meals program carried out on a large scale in Ghana demonstrated the positive impact of meals on children living below the poverty line (69). These results show that a school feeding program offering balanced meals throughout the school year helps to combat malnutrition among pupils. Good nutritional status and health during childhood and adolescence, in addition to education, are key factors in human capital (15). School feeding programs therefore contribute directly to human capital by improving the nutritional status of pupils and, more indirectly, their level of education. This impact can be measured conceptually by the Learning Adjusted Years of Schooling (LAYS) tool developed by the World Bank (70).

Studies have highlighted the negative effects of malnutrition and undernutrition on human capital. Malnourished children are more likely to miss out on school and to be less productive as adults, with incomes 10% lower over their lifetime (12, 14, 15). Poor nutritional status also leads to problems of underweight, thinness and stunted growth. Of these, stunting is a long-term problem that cannot be resolved by nutritional intake alone. The prevalence of stunting is also a good indicator of inequalities in human development (16, 17). Stunting in childhood leads to impaired brain development, lower cognitive skills and lower educational attainment, resulting in lower future earnings. It is often associated with negative lifelong consequences in terms of non-communicable diseases, leading to increased healthcare costs (12, 71). Stunting has a negative impact on cognitive abilities, education and skills development, jeopardising an economy’s labour productivity and human capital (17, 20) (5).

These results indicate that providing all primary schools with school canteen coverage over the entire school year would improve the nutritional status of pupils.

### 3.5. Complementary interventions

To increase the positive impact of school feeding programs on pupils, so-called complementary interventions must also be put in place for the benefit of pupils. Fortification of micronutrient foods, WASH (water, sanitation and hygiene) programs and the provision of deworming treatments can remedy micronutrient deficiencies, which are essential for children’s cognitive learning, and reduce school absenteeism due to illness (8, 12).

These interventions are termed complementary because they are not the only means of obtaining food, and because micronutrient supplementation and deworming can be carried out independently of school feeding (16).

Nevertheless, there is a strong case for integrating micronutrient fortification into school feeding programs, and also for deworming treatments to be carried out alongside school feeding programs wherever there is an epidemiological risk. Such interventions are now part of the policy of World Food Program (WFP) school feeding programs, and can therefore be considered indispensable (16). In the long term, school feeding programs, through their impact on human capital, enable the country to achieve a return on investment. A study carried out in Sweden shows that pupils who benefit during their entire primary school period have 3% greater lifetime income compared to unexposed pupils. Results shows that an additional year of school lunches increases adult income by 0.35% (64). The same study showed that the school feeding program didn’t lead to any major improvements in household income due to reduced food expenditures. In fact, the savings made on parents’ food expenditure appear too small to generate long-term effects on household income. These results don’t corroborate those of Dahl and Lochner, who show that school feeding programs can affect household income through reductions in food expenses and household work. This enables more parents to enter the labour market (72).

## Limitations and strengths

The methodology of this study has a number of limits. It was difficult to find in the economic literature practical methodologies for analysing the return on investment in school meals on the development of human capital. The main difficulty lies in calculating the transition probabilities from one state to another. This requires a great deal of statistical data, as well as estimates based on existing studies. Methodologies developed are very often theoretical and their application requires specific data that is rarely available. Consequently, results from studies carried out in other countries have been used. In the absence of data on the prevalence of stunting in school-age children, the prevalence in children under 5 in Burkina Faso was used (73). Also, the ratio of malnutrition rates between pupils who do not benefit from school canteens and those who do was calculated on the basis of the results of a study in Ghana (32) and Ethiopia (67) and the report of a review (74). With regard to data on education, certain values such as the school attendance ratio between malnourished and non-malnourished pupils and the mortality rate ratio between malnourished and non-malnourished children were selected on the basis of a subset of countries for which data was readily available. The nutritional quality of school meals was not taken into account in the study.

Since 2016, the security situation in Burkina Faso has also had a significant impact on the nutritional situation, and therefore on modeling. However, we took the view that by 2024, the base year for this simulation, the indicators used in this study already incorporate the security situation, and that consequently there was no longer any security scenario to develop.

The method enables an in-depth analysis of school feeding programs in a given country, and the transparency with which assumptions are formulated contributes to the relevance of the results. These results can be used as an advocacy tool by government stakeholders for greater investment in school feeding programs. A major advantage of this methodology is that it avoids double counting of beneficiaries.

## Conclusion

This study on the impact of the school feeding program on the development of human capital through the micro-simulation focuses on the effects of this program on the survival, nutrition and education of the beneficiary pupils. The results show that the school feeding program is a cost-effective investment in improving children’s health, nutrition and education, which are the main components of human capital development.

However, in Burkina Faso, the current level of coverage of this program does not allow school meals to be considered as an investment, but rather as expenditure.

The study concluded that in Burkina Faso, if school canteens are envisioned to be a means to contribute to the development of human capital, efforts should be redoubled to fill the funding gaps so that all schools are covered throughout the entire school year. The nutritional quality of the menus provided to pupils at mealtimes should also be improved and include a large quantity of foods rich in micronutrients and encourage the use of local foods as a way to promote a healthy and sustainable food system. School feeding programs are a sustainable way of investing in human capital.

## Author Contributions

D.S.O. and E.W.R.C. designed and carried out the study. G.A.N., T.S.Z.C. participated in data collection and curation, analysis and interpretation. D.S.O. writing and editing the manuscript. J.D., O.O., S.D. and I.K. made critical revisions to the article. The final manuscript was approved by M.H.D. All authors have read and approved the final manuscript as submitted.

## Funding

This research received no funding.

## Institutional Review Board Statement

This study was conducted according to the guidelines laid down in the Declaration of Helsinki and all procedures involving human subjects were approved by the Health Research Ethics Committee (CERS), in deliberation No. 2021-11-267 of Burkina Faso on 14 February 2022; under the ethics approval 2022_33_/MS/MESRSI/CERS.

## Informed Consent Statement

Written informed consent has been obtained from the patient(s) to publish this paper.

## Data Availability Statement

The raw data supporting the conclusions of this article will be made available by the authors on request.

## Declaration of conflicting interests

The authors declare no conflicts of interest related to this study.

## Acknowledgements

We would like to thank the Regional Department of Pre-school, Primary and Non-Formal Education and the teaching staff of the elementary school in the Plateau Central region. We would also like to thank the schoolchildren who took part in the study, as well as the data collection team.

## Notes

### Competing Interest Statement

The authors have declared no competing interest.

### Author Declarations

The study was officially approved by the Burkina Faso Health Research Ethics Committee (CERS) with reference N 2022_33_/MS/MESRSI/CERS of 14-02-202.

